# Modifiable and non-modifiable risk factors for COVID-19: results from UK Biobank

**DOI:** 10.1101/2020.04.28.20083295

**Authors:** Frederick K Ho, Carlos A Celis-Morales, Stuart R Gray, S Vittal Katikireddi, Claire L Niedzwiedz, Claire Hastie, Donald M Lyall, Lyn D Ferguson, Colin Berry, Daniel F Mackay, Jason MR Gill, Jill P Pell, Naveed Sattar, Paul Welsh

**Affiliations:** Institute of Health and Wellbeing, University of Glasgow, 1 Lilybank Gardens, Glasgow, G12 8RZ, United Kingdom; Institute of Cardiovascular and Medical Sciences, University of Glasgow, BHF Glasgow Cardiovascular Research Centre, 126 University Place, Glasgow, G12 8TA, United Kingdom

**Author notes:** Joint first authors. Joint senior. **Address for correspondence** Paul Welsh OR Naveed Sattar, Institute of Cardiovascular and Medical Sciences, University of Glasgow, BHF Glasgow Cardiovascular Research Centre, 126 University Place, Glasgow G12 8TA, UK.

## Abstract

**Background:** Information on risk factors for COVID-19 is sub-optimal. We investigated demographic, lifestyle, socioeconomic, and clinical risk factors, and compared them to risk factors for pneumonia and influenza in UK Biobank.

**Methods:** UK Biobank recruited 37–70 year olds in 2006–2010 from the general population. The outcome of confirmed COVID-19 infection (positive SARS-CoV-2 test) was linked to baseline UK Biobank data. Incident influenza and pneumonia were obtained from primary care data. Poisson regression was used to study the association of exposure variables with outcomes.

**Findings:** Among 428,225 participants, 340 had confirmed COVID-19. After multivariable adjustment, modifiable risk factors were higher body mass index (RR 1.24 per SD increase), smoking (RR 1.38), slow walking pace as a proxy for physical fitness (RR 1.66) and use of blood pressure medications as a proxy for hypertension (RR 1.40). Non-modifiable risk factors included older age (RR 1.10 per 5 years), male sex (RR 1.64), black ethnicity (RR 1.86), socioeconomic deprivation (RR 1.26 per SD increase in Townsend Index), longstanding illness (RR 1.38) and high cystatin C (RR 1.24 per 1 SD increase). The risk factors overlapped with pneumonia somewhat; less so for influenza. The associations with modifiable risk factors were generally stronger for COVID-19, than pneumonia or influenza.

**Interpretation:** These findings suggest that modification of lifestyle may help to reduce the risk of COVID-19 and could be a useful adjunct to other interventions, such as social distancing and shielding of high risk.

**Funding:** British Heart Foundation, Medical Research Council, Chief Scientist Office.

## Introduction

COVID-19, caused by SARS-CoV-2 infection, includes a spectrum of morbidity from asymptomatic infection ^1^ to severe pneumonia in patients presenting for medical care ^2^. The COVID-19 pandemic ^3^ has led to concerted research efforts to identify people at greatest risk of developing the infection and progressing to critical illness and dying. Predictors of disease severity include older age, smoking, diabetes, hypertension, kidney disease, COPD, and previous cardiovascular disease ^4–7^. In addition it is becoming clear that other risk factors might include obesity and low physical fitness ^8,9^ However, many studies investigate risk factors for disease progression to death or critical illness among hospitalised patients^7^ as opposed to healthy comparators.

Identifying the risk factors for COVID-19 is important in terms of identifying factors that can be modified to reduce risk, as well as identifying non-modifiable risk factors that can help identify high risk groups who require shielding and targeting for testing, and eventual vaccine and anti-viral therapies. Pneumonia is a life-threatening complication of COVID-19 infection ^10^. Established major risk factors for community-acquired pneumonia include many of the emerging risk factors for COVID-19 ^11^. As COVID-19 is caused by SARS-CoV-2 viral infection, it may have risk factors for contraction of the disease that are common to other respiratory virus conditions such as Influenza.

UK Biobank is a large prospective, deeply-phenotyped, population-based cohort study carried out in the UK ^12^. Over the study period, testing for COVID-19 in England was conducted in A&E departments and in-hospital. These data were provided by Public Health England (PHE) and linked to UK Biobank baseline data. We aimed to establish modifiable and non-modifiable risk factors for confirmed COVID-19. We also aimed to compare these risk factors to risk factors for the incident pneumonia over a similar time span.

## Methods

UK Biobank was conducted via 22 assessment centres across England, Scotland, and Wales between March 2006 and December 2010 and recruited 502,624 participants aged 37 to 73 years. The present study was restricted to participants living in England for whom COVID-19 test results were available. Death data were available to the end of January 2018 in England. To reduce bias, we excluded from the study all participants known to have died COVID-19 pandemic. Baseline biological measurements were recorded and touch-screen questionnaires were administered according to a standardised protocol. ^12,13^ UK Biobank received ethical approval from the North West Multi-Centre Research Ethics Committee (REC reference: 11/NW/03820). All participants gave written informed consent before enrolment in the study, which was conducted in accord with the principles of the Declaration of Helsinki. This study was performed under UK Biobank application number 7155.

### Outcomes

Results of COVID-19 tests for UK Biobank participants were provided by PHE ^7^. Data provided by PHE included the specimen date, specimen type (e.g. upper respiratory tract), laboratory, origin (whether evidence from microbiological record that the participant was an inpatient or not) and result (positive or negative). Data were available for the period 16th March 2020 to 14th April 2020. Results were available for 2,724 tests conducted on 1,474 individuals. Confirmed COVID-19 infection (primary outcome) was defined as at least one positive result in the context of an in-hospital or accident and emergency (A&E) test. Any positive test result was used as a sensitivity analysis.

Pneumonia was defined based on ICD-10 codes J12-J18, and influenza based on J09-J11, converted into Read Codes using the UK Biobank’s look-up table. Incident pneumonia and influenza, occurring after 1^st^ January 2016 (an arbitrary date taken to mimic the time lag from baseline exposure measurement to incident COVID-19 infection, while also obtaining sufficient case numbers) was obtained from a 41% sample of participants with available data from primary care. A sensitivity analysis was conducted using cases after 1^st^ January 2015 and demonstrated consistent data. An analysis was also conducted exploring risk factors for all cases of pneumonia and influenza after baseline.

### Exposures

Exposures were measured at the baseline assessment visit between 2006 and 2010. Ethnicity, smoking, alcohol consumption, physician-diagnosed prevalent conditions and medication use were self-reported. For the present analyses, ethnicity was coded as white, south Asian, black, or mixed/other. Smoking status was categorised into never versus former/current smoking. Systolic and diastolic blood pressures were measured at the baseline visit, preferentially using an automated measurement, but using manual measurement where this was not available, and average of available measures used. Area-level socioeconomic deprivation was assessed by the Townsend score (incorporating measures of unemployment, non-car ownership, non-home ownership and household overcrowding) corresponding to the participants’ home postcode. Higher scores on the Townsend score represent greater socioeconomic deprivation. Self-reported walking pace was rated by each participant as slow, steady/average, or brisk ^14^.

The definition of baseline diabetes included self-reported type 1 or type 2 diabetes, those with a primary or secondary hospital diagnoses relating to diabetes at baseline (ICD-10 codes E10-E14.9), and those who reported using diabetes medications. Baseline cardiovascular disease was defined as self-reported myocardial infarction, stroke, or transient ischaemic attack. Cancer, longstanding illness, and depression were self-reported on touchscreen questionnaire. Other previous health complaints including asthma, rheumatoid arthritis, CKD, systemic lupus erythematosus (SLE), sleep apnoea, COPD, pneumonia, bronchitis (including bronchitis, bronchiectasis, and emphysema), and other respiratory diseases (including interstitial lung disease, asbestosis, pulmonary fibrosis, alveolitis, respiratory failure, pleurisy, pneumothorax, other respiratory condition) which were derived from self-report at nurse interview. Some conditions did not have sufficient numbers of cases for multivariable analysis, but univariable results are presented for completeness. Statin (categorised to include other cholesterol lowering medications) and blood pressure medication use were also recorded from self-report, with blood pressure medication being used as a proxy for baseline diagnosed hypertension.

BMI is the ratio of the measured body mass in kg divided by height squared measured in metres. Height was measured using a Seca 202 height measure. Weight and whole-body fat mass and fat free mass were measured to the nearest 0.1 kg using the Tanita BC-418 MA body composition analyser. Socks and shoes were removed when height was measured Grip strength was measured using a Jamar J00105 hydraulic hand dynamometer and the mean was derived from the right and left hand values expressed in kilograms ^15^.

Lung function was assessed by spirometry using a Vitalograph Pneumotrac 6800 spirometer (Vitalograph, Buckingham, UK). Participants did not perform spirometry if they answered yes to unsure to the following: chest infection in the last month, history of detached retina, heart attack, surgery to eyes, chest or abdomen in last 3 months, history of collapsed lung, pregnancy, or currently on medication for tuberculosis. The aim was to record two acceptable blows from a maximum of three attempts. The spirometer software compared the acceptability of the first two blows and, if acceptable (defined as a ≤5% difference in FVC and FEV1), the third blow was not required. The mean observation was taken for both measures.

Blood collection sampling procedures for the study have been previously described and validated.^16^ Biochemical assays were performed at a dedicated central laboratory on around 480,000 samples. Further details of these measurements can be found in the UK Biobank Data Showcase and Protocol (http://www.ukbiobank.ac.uk). For the present study we included total cholesterol, HDL cholesterol, rheumatoid factor, cystatin C, glycated haemoglobin (HbA1C), C-reactive protein (CRP), differential white blood cell count, and red cell distribution width as exposures of interest.

### Statistics

Mean and standard deviations were reported for continuous outcomes except for biomarkers, where median and interquartile ranges (IQR) were reported. Poisson regression with robust ‘sandwich’ standard errors were used to study the associations of exposure variables with confirmed COVID-19 and pneumonia. Poisson regressions were used because they provide risk ratio (RR) which is easier to interpret and robust error estimation ensures accurate inference ^17^. Three adjustment schemes were considered: model 0 - univariate (i.e. no adjustment), model 1 – adjusted for age, sex, ethnicity, and deprivation index, and model 2 – further adjusted for behavioural (smoking and alcohol drinking) and physical (adiposity, blood pressure, spirometry, and physical capability) factors that were found to be significant in model 1. To avoid multicollinearity, if there were similar factors that were significant in model 2, only the one with strongest association was chosen. This included, for example, choosing one from, BMI, BMI categories, body fat-free mass, body fat mass, and body fat percent, or one from FEV1, FVC, and FEV1/FVC. For continuous variables, the linearity of exposure-outcome associations were tested using penalised cubic splines in generalised additive model (GAM) ^18^. Nonlinearity was tested using likelihood ratio test comparing a model with the exposure fitted on a spline with a model assuming a linear exposure-outcome relationship. P-value for nonlinearity < 0.05 suggest evidence against the linearity assumption. Spline smoothness was chosen using generalised cross validation ^19^. Population attributable fractions (PAFs) were calculated to determine the relative contribution of each risk factor to the overall number of confirmed COVID-19 cases within UK Biobank. Another Poisson model which included all significant factors in model 2 was fitted to estimate the mutually adjusted RRs. These RRs were biased towards null because of over adjustment bias but were used to construct the PAFs to ensure the PAF estimates did not overlap or exceed 100%. In general, two-tailed P-values < 0.05 were considered statistically significant. Analyses were conducted in R Statistical Software version 3.5.3 with the package “mgcv”.

## Results

Of 445,857 participants in England, 428,225 were alive during the available follow-up period. Complete data on covariates were available for 285,817 (66.7%) participants. Primary care data on incident pneumonia were available in 117,948 (41.3%) participants (Supplementary Fig 1). At 1^st^ March 2020, the age range of eligible participants was 48-85 years, and time elapsed from baseline was median 10.97 years (IQR 10.36-11.55 years). Of these participants, 898 received at least one COVID-19 test, and 400 had at least one positive result, with 340 positive results conducted in hospital or A&E (primary outcome).

### Univariable risk factors for incident COVID-19

Key univariable potentially modifiable risk factors for confirmed COVID-19 included current and former smoking (RR 1.62), higher BMI and body fat (RR 2.29 for obesity), treated hypertension (RR 2.07), higher systolic and diastolic blood pressure, and slow walking pace as a proxy for fitness (RR 2.40) (Table 1). Among non-modifiable risk factors were older age (particularly the over 65s at baseline, corresponding to over 75 in 2020), male sex (RR 1.56), black ethnicity (RR 2.74), socioeconomic deprivation (RR 1.37 per 1 SD increase in Townsend Index) (Table 1). COVID-19 was only weakly associated with low FEV1/FVC ratio (RR 0.90 per 1 SD increase). Among baseline comorbidities, general long standing illness (RR 1.88), baseline diabetes (RR 2.07), baseline CVD (RR 2.19), sleep apnoea (RR 3.91), statin use (RR 1.85), higher inflammatory markers (particularly white blood cell count; RR 1.20 per 1 SD increase), and higher cystatin C (RR 1.45 per 1 SD increase) were also associated with confirmed COVID-19 (Table 1). Risk factor associations were similar when all 400 test positive COVID-19 cases were considered, rather than only in-hospital positive tests (supplementary table 1)

**Table 1.** Univariable association of baseline risk factors with COVID-19 in 2020, pneumonia occurring after 2016, and influenza occurring after 2016

|  | Covid-19 |  |  |  | Pneumonia (after 2016) |  |  |  | Influenza (after 2016) |  |  |  |
| --- | --- | --- | --- | --- | --- | --- | --- | --- | --- | --- | --- | --- |
|  | No<br>n=285477 | Yes<br>n=340 | RR* | P | No<br>n=117689 | Yes<br>n=259 | RR | P | No<br>n=117837 | Yes<br>n=111 | RR | P |
| Age (years) | 56.15 (8.12) | 57.66 (8.49) | 1.21† | 0.0006 | 56.23 (8.09) | 58.29 (7.57) | 1.31 | < 0.0001 | 56.24 (8.09) | 56.11 (7.99) | 0.98 | 0.87 |
| Age categories (years), n (%) |  |  |  | < 0.0001 |  |  |  | 0.0005 |  |  |  | 0.41 |
| <50 | 71218 (24.95) | 76 (22.35) | 1 | REF | 28967 (24.61) | 40 (15.44) | 1 | REF | 28983 (24.60) | 24 (21.62) | 1 | REF |
| 50-54 | 44697 (15.66) | 41 (12.06) | 0.86 | 0.44 | 18257 (15.51) | 29 (11.20) | 1.15 | 0.57 | 18262 (15.50) | 24 (21.62) | 1.59 | 0.11 |
| 55-59 | 51240 (17.95) | 51 (15.00) | 0.93 | 0.70 | 21184 (18.00) | 58 (22.39) | 1.98 | 0.0009 | 21220 (18.01) | 22 (19.82) | 1.25 | 0.45 |
| 60-64 | 67559 (23.67) | 78 (22.94) | 1.08 | 0.63 | 28256 (24.01) | 79 (30.50) | 2.02 | 0.0003 | 28312 (24.03) | 23 (20.72) | 0.98 | 0.95 |
| ≥65 | 50763 (17.78) | 94 (27.65) | 1.73 | 0.0004 | 21025 (17.86) | 53 (20.46) | 1.82 | 0.004 | 21060 (17.87) | 18 (16.22) | 1.03 | 0.92 |
| Male sex, n (%) | 131395 (46.03) | 194 (57.06) | 1.56 | < 0.0001 | 54098 (45.97) | 88 (33.98) | 0.61 | 0.0001 | 54134 (45.94) | 52 (46.85) | 1.04 | 0.85 |
| Ethnicity (%) |  |  |  | 0.0004 |  |  |  | 0.54 |  |  |  | 0.93 |
| White | 270360 (94.70) | 307 (90.29) | 1 | REF | 112010 (95.17) | 248 (95.75) | 1 | REF | 112153 (95.18) | 105 (94.59) | 1 | REF |
| Black | 4163 (1.46) | 13 (3.82) | 2.74 | 0.0004 | 1266 (1.08) | 4 (1.54) | 1.43 | 0.48 | 1269 (1.08) | 1 (0.90) | 0.84 | 0.86 |
| South Asian | 4701 (1.65) | 7 (2.06) | 1.31 | 0.48 | 2225 (1.89) | 5 (1.93) | 1.01 | 0.97 | 2228 (1.89) | 2 (1.80) | 0.96 | 0.95 |
| Others | 6253 (2.19) | 13 (3.82) | 1.83 | 0.03 | 2188 (1.86) | 2 (0.77) | 0.41 | 0.21 | 2187 (1.86) | 3 (2.70) | 1.46 | 0.51 |
| Deprivation index (score) | -1.40 (3.01) | -0.31 (3.41) | 1.37 | < 0.0001 | -1.44 (2.93) | -1.11 (3.15) | 1.12 | 0.07 | -1.44 (2.93) | -0.85 (3.30) | 1.21 | 0.03 |
| Current/former smoker, n (%) | 126186 (44.20) | 191 (56.18) | 1.62 | < 0.0001 | 52058 (44.23) | 120 (46.33) | 1.09 | 0.50 | 52126 (44.24) | 52 (46.85) | 1.11 | 0.58 |
| Alcohol drinking status, n (%) |  |  |  | 0.001 |  |  |  | 0.03 |  |  |  | 0.77 |
| Never | 11983 (4.20) | 19 (5.59) | 1 | REF | 5033 (4.28) | 17 (6.56) | 1 | REF | 5045 (4.28) | 5 (4.50) | 1 | REF |
| Former | 9259 (3.24) | 22 (6.47) | 1.5 | 0.20 | 3893 (3.31) | 14 (5.41) | 1.06 | 0.86 | 3902 (3.31) | 5 (4.50) | 1.29 | 0.68 |
| Current | 264235 (92.56) | 299 (87.94) | 0.71 | 0.15 | 108763 (92.42) | 228 (88.03) | 0.62 | 0.06 | 108890 (92.41) | 101 (90.99) | 0.94 | 0.89 |
| BMI (kg/m²) | 27.27 (4.60) | 29.02 (5.25) | 1.38 | < 0.0001 | 27.38 (4.68) | 28.43 (5.40) | 1.23 | 0.0003 | 27.39 (4.68) | 27.72 (5.00) | 1.07 | 0.49 |
| BMI categories, n (%) |  |  |  | < 0.0001 |  |  |  | 0.16 |  |  |  | 0.18 |
| Underweight | 1245 (0.44) | 1 (0.29) | 1.05 | 0.96 | 490 (0.42) | 1 (0.39) | 1.01 | 0.99 | 491 (0.42) | 0 (0.00) | 0.00 | 0.98 |
| Normal | 95864 (33.58) | 73 (21.47) | 1 | REF | 38492 (32.71) | 78 (30.12) | 1 | REF | 38529 (32.70) | 41 (36.94) | 1 | REF |
| Overweight | 121857 (42.69) | 150 (44.12) | 1.62 | 0.0008 | 50404 (42.83) | 102 (39.38) | 1.00 | 0.99 | 50469 (42.83) | 37 (33.33) | 0.69 | 0.10 |
| Obese | 66511 (23.30) | 116 (34.12) | 2.29 | < 0.0001 | 28303 (24.05) | 78 (30.12) | 1.36 | 0.06 | 28348 (24.06) | 33 (29.73) | 1.09 | 0.70 |
| Body fat-free mass (Kg) | 53.44 (11.46) | 56.26 (11.40) | 1.26 | < 0.0001 | 53.44 (11.55) | 51.62 (12.00) | 0.85 | 0.01 | 53.44 (11.55) | 53.56 (11.98) | 1.01 | 0.88 |
| Body fat mass (Kg) | 24.48 (9.23) | 27.21 (10.37) | 1.3 | < 0.0001 | 24.70 (9.43) | 27.51 (10.96) | 1.30† | < 0.0001 | 24.71 (9.43) | 24.93 (10.27) | 1.03 | 0.78 |
| Body fat proportion (percent) | 31.10 (8.46) | 32.21 (8.56) | 1.14 | 0.02 | 31.29 (8.49) | 34.27 (8.76) | 1.41 | < 0.0001 | 31.29 (8.49) | 31.27 (9.03) | 1.00 | 0.98 |
| Systolic blood pressure (mmHg) | 137.15 (18.14) | 139.27 (18.57) | 1.12 | 0.03 | 137.47 (18.38) | 137.31 (17.63) | 0.99 | 0.90 | 137.47 (18.38) | 137.30 (18.41) | 0.99 | 0.93 |
| Diastolic blood pressure (mmHg) | 82.01 (9.92) | 83.53 (10.46) | 1.16 | 0.005 | 82.13 (10.03) | 81.15 (9.70) | 0.91 | 0.12 | 82.13 (10.03) | 82.36 (10.92) | 1.03 | 0.79 |
| FEV1(litres) | 2.75 (0.77) | 2.66 (0.73) | 0.89 | 0.03 | 2.75 (0.79) | 2.42 (0.81) | 0.64† | < 0.0001 | 2.75 (0.79) | 2.66 (0.86) | 0.89 | 0.21 |
| FVC (litres) | 3.64 (0.98) | 3.56 (0.93) | 0.92 | 0.15 | 3.64 (1.04) | 3.26 (1.01) | 0.66† | < 0.0001 | 3.64 (1.04) | 3.52 (1.03) | 0.89 | 0.22 |
| FEV1/FVC | 0.76 (0.07) | 0.75 (0.08) | 0.9 | 0.04 | 0.76 (0.07) | 0.74 (0.10) | 0.84† | 0.002 | 0.76 (0.07) | 0.75 (0.08) | 0.94 | 0.49 |
| Walking pace (%) |  |  |  | < 0.0001 |  |  |  | < 0.0001 |  |  |  | 0.65 |
| Slow | 19101 (6.69) | 54 (15.88) | 2.40 | < 0.0001 | 8026 (6.82) | 35 (13.51) | 1.90 | 0.0006 | 8051 (6.83) | 10 (9.01) | 1.33 | 0.40 |
| Average | 150594 (52.75) | 177 (52.06) | 1 | REF | 62121 (52.78) | 142 (54.83) | 1 | REF | 62205 (52.79) | 58 (52.25) | 1 | REF |
| Brisk | 115782 (40.56) | 109 (32.06) | 0.80 | 0.07 | 47542 (40.40) | 82 (31.66) | 0.75 | 0.04 | 47581 (40.38) | 43 (38.74) | 0.97 | 0.88 |
| Grip strength (Kg) | 30.85 (10.93) | 31.21 (10.77) | 1.03† | 0.55 | 30.74 (11.04) | 27.68 (10.77) | 0.75 | < 0.0001 | 30.74 (11.04) | 30.62 (11.80) | 0.98 | 0.87 |
| Prevalent disease at baseline (%) |  |  |  |  |  |  |  |  |  |  |  |  |
| Longstanding illness, n (%) | 83006 (29.08) | 148 (43.53) | 1.88 | < 0.0001 | 35361 (30.05) | 108 (41.70) | 1.66 | < 0.0001 | 35427 (30.06) | 42 (37.84) | 1.42 | 0.08 |
| Diabetes | 13626 ( 4.77) | 32 ( 9.41) | 2.07 | < 0.0001 | 5665 ( 4.81) | 25 ( 9.65) | 2.11 | 0.0004 | 5684 ( 4.82) | 6 ( 5.41) | 1.13 | 0.78 |
| CVD | 13785 ( 4.83) | 34 (10.00) | 2.19 | < 0.0001 | 5977 ( 5.08) | 21 ( 8.11) | 1.65 | 0.03 | 5994 ( 5.09) | 4 ( 3.60) | 0.70 | 0.48 |
| Cancer | 19368 ( 6.80) | 31 ( 9.25) | 1.4 | 0.08 | 8034 ( 6.85) | 30 (11.58) | 1.78 | 0.003 | 8052 ( 6.85) | 12 (10.81) | 1.65 | 0.10 |
| Depression | 54199 (18.99) | 62 (18.24) | 0.95 | 0.72 | 22173 (18.84) | 36 (13.90) | 0.70 | 0.04 | 22188 (18.83) | 21 (18.92) | 1.01 | 0.98 |
| CKD stages 3-5 | 353 ( 0.12) | 2 ( 0.59) | 4.76 | 0.03 | 152 ( 0.13) | 0 ( 0.00) | 0.00 | 0.97 | 152 ( 0.13) | 0 ( 0.00) | 0.00 | 0.98 |
| SLE | 341 ( 0.12) | 0 ( 0.00) | 0 | 0.96 | 151 ( 0.13) | 0 ( 0.00) | 0.00 | 0.97 | 151 ( 0.13) | 0 ( 0.00) | 0.00 | 0.98 |
| Asthma | 31367 (10.99) | 47 (13.82) | 1.3 | 0.10 | 12986 (11.03) | 38 (14.67) | 1.39 | 0.06 | 13010 (11.04) | 14 (12.61) | 1.16 | 0.60 |
| Sleep apnoea | 864 ( 0.30) | 4 ( 1.18) | 3.91 | 0.007 | 383 ( 0.33) | 0 ( 0.00) | 0.00 | 0.97 | 383 ( 0.33) | 0 ( 0.00) | 0.00 | 0.97 |
| COPD | 608 ( 0.21) | 2 ( 0.59) | 2.77 | 0.15 | 271 ( 0.23) | 1 ( 0.39) | 1.68 | 0.61 | 271 ( 0.23) | 1 ( 0.90) | 3.93 | 0.17 |
| Bronchitis | 2920 ( 1.02) | 8 ( 2.35) | 2.33 | 0.02 | 1275 ( 1.08) | 6 ( 2.32) | 2.16 | 0.06 | 1277 ( 1.08) | 4 ( 3.60) | 3.40 | 0.02 |
| Pneumonia | 3535 ( 1.24) | 5 ( 1.47) | 1.19 | 0.70 | 1384 ( 1.18) | 5 ( 1.93) | 1.65 | 0.27 | 1388 ( 1.18) | 1 ( 0.90) | 0.76 | 0.79 |
| Other respiratory disease | 1488 ( 0.52) | 3 ( 0.88) | 1.7 | 0.36 | 609 ( 0.52) | 1 ( 0.39) | 0.75 | 0.77 | 610 ( 0.52) | 0 ( 0.00) | 0.00 | 0.97 |
| Medication at baseline, n (%) |  |  |  |  |  |  |  |  |  |  |  |  |
| Statin | 44164 (15.47) | 86 (25.29) | 1.85 | < 0.0001 | 18733 (15.92) | 47 (18.15) | 1.17 | 0.33 | 18770 (15.93) | 10 ( 9.01) | 0.52 | 0.050 |
| BP medication | 45493 (15.94) | 96 (28.24) | 2.07 | < 0.0001 | 19078 (16.21) | 42 (16.22) | 1.00 | 1.00 | 19106 (16.21) | 14 (12.61) | 0.75 | 0.31 |
| Steroid | 1070 ( 0.37) | 2 ( 0.59) | 1.57 | 0.52 | 440 ( 0.37) | 2 ( 0.77) | 2.07 | 0.31 | 442 ( 0.38) | 0 ( 0.00) | 0.00 | 0.98 |
| Biomarker at baseline |  |  |  |  |  |  |  |  |  |  |  |  |
| Total cholesterol (mmol/L) | 5.62 (4.91-6.33) | 5.44 (4.62-6.20) | 0.83† | 0.0004 | 5.62 (4.90-6.34) | 5.62 (4.84-6.28) | 0.99 | 0.85 | 5.62 (4.90-6.34) | 5.70 (4.96-6.29) | 1.07 | 0.48 |
| HDL cholesterol (mmol/L) | 1.40 (1.18-1.68) | 1.29 (1.07-1.53) | 0.70 | < 0.0001 | 1.40 (1.17-1.67) | 1.41 (1.16-1.70) | 1.00 | 0.94 | 1.40 (1.17-1.67) | 1.36 (1.11-1.62) | 0.87 | 0.15 |
| Cystatin C (mg/L) | 0.88 (0.80-0.97) | 0.94 (0.85-1.04) | 1.45 | < 0.0001 | 0.88 (0.80-0.98) | 0.92 (0.81-1.01) | 1.20 | 0.001 | 0.88 (0.80-0.98) | 0.91 (0.82-0.99) | 1.09 | 0.36 |
| HbA1c (mmol/mol) | 35.10 (32.60-37.70) | 36.30 (33.40-39.70) | 1.28† | < 0.0001 | 35.10 (32.60-37.70) | 36.10 (33.70-38.95) | 1.20 | < 0.0001 | 35.10 (32.60-37.70) | 35.20 (32.40-38.25) | 1.10 | 0.22 |
| CRP (mg/L) | 1.26 (0.63-2.58) | 1.68 (0.88-3.12) | 1.15† | 0.0006 | 1.27 (0.63-2.61) | 1.51 (0.91-3.17) | 1.14 | 0.003 | 1.27 (0.64-2.61) | 1.46 (0.77-3.06) | 1.12 | 0.12 |
| Rheumatoid factor (IU/ml) | 3.16 (3.16-3.16) | 3.16 (3.16-3.16) | 0.98 | 0.73 | 3.16 (3.16-3.16) | 3.16 (3.16-3.16) | 1.04 | 0.46 | 3.16 (3.16-3.16) | 3.16 (3.16-3.16) | 1.04 | 0.64 |
| Red cell distribution width (%) | 13.31 (12.90-13.81) | 13.32 (12.98-13.95) | 1.12 | 0.02 | 13.30 (12.89-13.80) | 13.40 (12.96-14.07) | 1.15 | 0.010 | 13.30 (12.89-13.80) | 13.29 (12.98-13.79) | 0.97 | 0.73 |
| White cell count (10<sup>9</sup> cells/Litre) | 6.60 (5.61-7.80) | 6.85 (5.76-8.27) | 1.20 | 0.0004 | 6.64 (5.65-7.81) | 7.02 (5.78-8.42) | 1.23 | 0.0004 | 6.64 (5.65-7.82) | 6.60 (5.51-7.86) | 1.04 | 0.64 |
| Neutrophil count (10<sup>9</sup> cells/Litre) | 4.00 (3.25-4.90) | 4.12 (3.25-5.10) | 1.11 | 0.051 | 4.01 (3.27-4.93) | 4.18 (3.39-5.27) | 1.14 | 0.02 | 4.01 (3.27-4.93) | 3.98 (3.26-5.09) | 1.01 | 0.92 |
| Lymphocyte count (10<sup>9</sup> cells/Litre) | 1.88 (1.52-2.29) | 1.94 (1.55-2.43) | 1.19† | 0.0008 | 1.88 (1.52-2.30) | 1.98 (1.62-2.44) | 1.20 | 0.002 | 1.88 (1.52-2.30) | 1.90 (1.54-2.52) | 1.07† | 0.47 |
| Monocyte count (10<sup>9</sup> cells/Litre) | 0.45 (0.36-0.56) | 0.48 (0.38-0.60) | 1.17 | 0.002 | 0.45 (0.36-0.56) | 0.48 (0.38-0.60) | 1.12 | 0.06 | 0.45 (0.36-0.57) | 0.49 (0.40-0.57) | 1.11† | 0.27 |

Numbers represent number (%) for categorical variables, mean (SD) for gaussian continuous variables, and median (25-75<sup>th</sup> percentiles) for skewed variables
\*Univariable relative risk ratio per 1 standard deviation for continuous variables and using comparator group for categorical variables.
†Evidence for non-linearity (see supplementary material)

### Univariable risk factors for incident pneumonia

Of the 117,837 participants with primary care data, 259 had pneumonia recorded after 2016. Of the modifiable risk factors pneumonia was associated with BMI and slow walking pace, but not smoking, blood pressure or blood pressure medication use. Incident pneumonia was more common in participants who were over 55 years at baseline, women (RR 0.61 among men), and socioeconomically deprived participants (RR 1.37 per 1 SD increase in Townsend Index), (Table 1). Pneumonia was also more common in those with low grip strength, and lower FEV and FVC, and FEV/FVC ratio (Table 1). Among comorbidities, baseline diabetes, CVD and cancer were approximately twice as common in those who developed pneumonia (Table 1), and any longstanding illness was also strong associated with pneumonia. Among blood biomarkers, poorer renal function as measured by higher cystatin-C, higher HbA1C, higher CRP and white blood cell count (including neutrophils, monocytes, and lymphocytes) and higher red cell distribution width were all associated with pneumonia. Ethnicity was not associated with pneumonia.

When investigating risk factors for pneumonia over the full follow-up time from baseline, risk factors were generally similar although there was increased power due to a higher number of incident cases (Supplementary table 1). In this analysis smoking was weakly associated with pneumonia (RR 1.17) as was blood pressure medication use (RR 1.15). Similar findings were found when we studied pneumonia occurring after 2015 (Supplementary Table 2).

### Univariable risk factors for incident influenza

Of the 117,837 participants with primary care data, 111 had influenza recorded after 2016. Those who developed influenza were generally more socioeconomically deprived, more likely to have bronchitis at baseline, and less likely to take statins. No other risk factors showed strong associations (Table 1). When all influenza cases occurring after baseline were considered, evidence of statistically significant associations became stronger due to increased power but cases were more common in slightly younger people and in women, and smoking, walking pace, and BMI was less strongly associated with influenza cases than for COVID-19, and blood pressure medication was not at all associated with influenza (Supplementary Table 1).

### Multivariable models for COVID-19 and pneumonia

After multivariable adjustment for sociodemographic factors, the modifiable risk factors for confirmed COVID-19 included smoking (RR 1.44 (95% CI 1.15-1.79)), higher BMI (RR 1.33 per SD increase (95% CI 1.21-1.46)) and other measures of body fat, diastolic blood pressure, as well as blood pressure medication, FEV1 and FVC (but not FEV1/FVC ratio), and slow walking pace (RR 2.04 (95% CI 1.49-2.79)) (Table 2). Other risk factors were older age (RR 1.15 per 5 years (95% CI 1.08-1.24)), male sex (RR 1.54 (95% CI 1.24-1.91)), black ethnicity (RR 2.07 (95% CI 1.16, 3.71)), socioeconomic deprivation (RR 1.35 per SD increase in Townsend Index (95% CI 1.23-1.49)). Among comorbidities, risk factors included longstanding illness (RR 1.65 (95% CI 1.32-2.06)), diabetes, CVD and statin use. Among blood biomarkers, risk factors included lower total and HDL-cholesterol, and higher cystatin C (RR 1.36 per 1 SD increase (95% CI 1.23-1.49)), HbA1C (RR 1.20 per 1 SD increase (95% CI 1.11-1.30)), CRP and white blood cell count (Table 2).

**Table 2.**
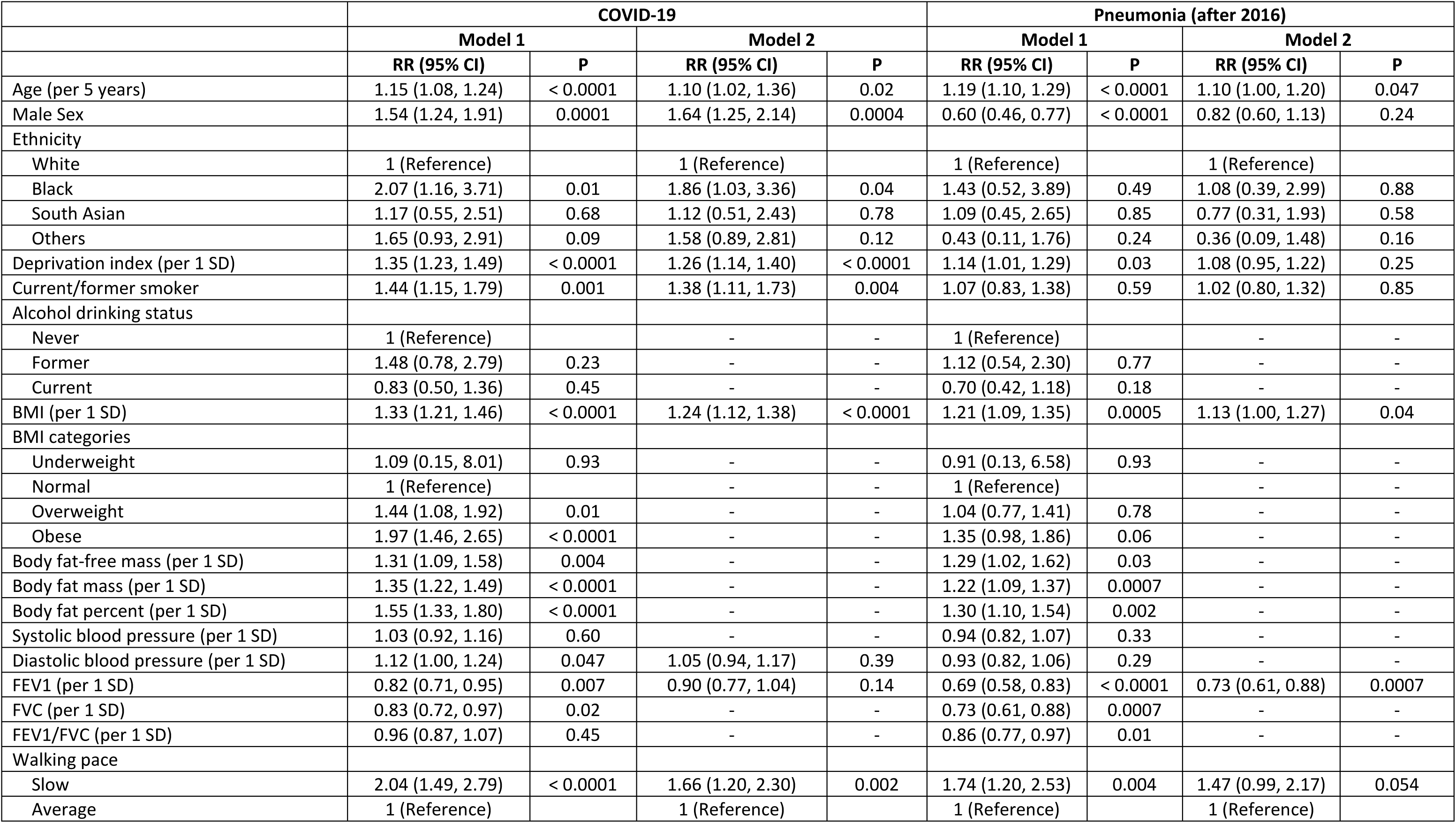
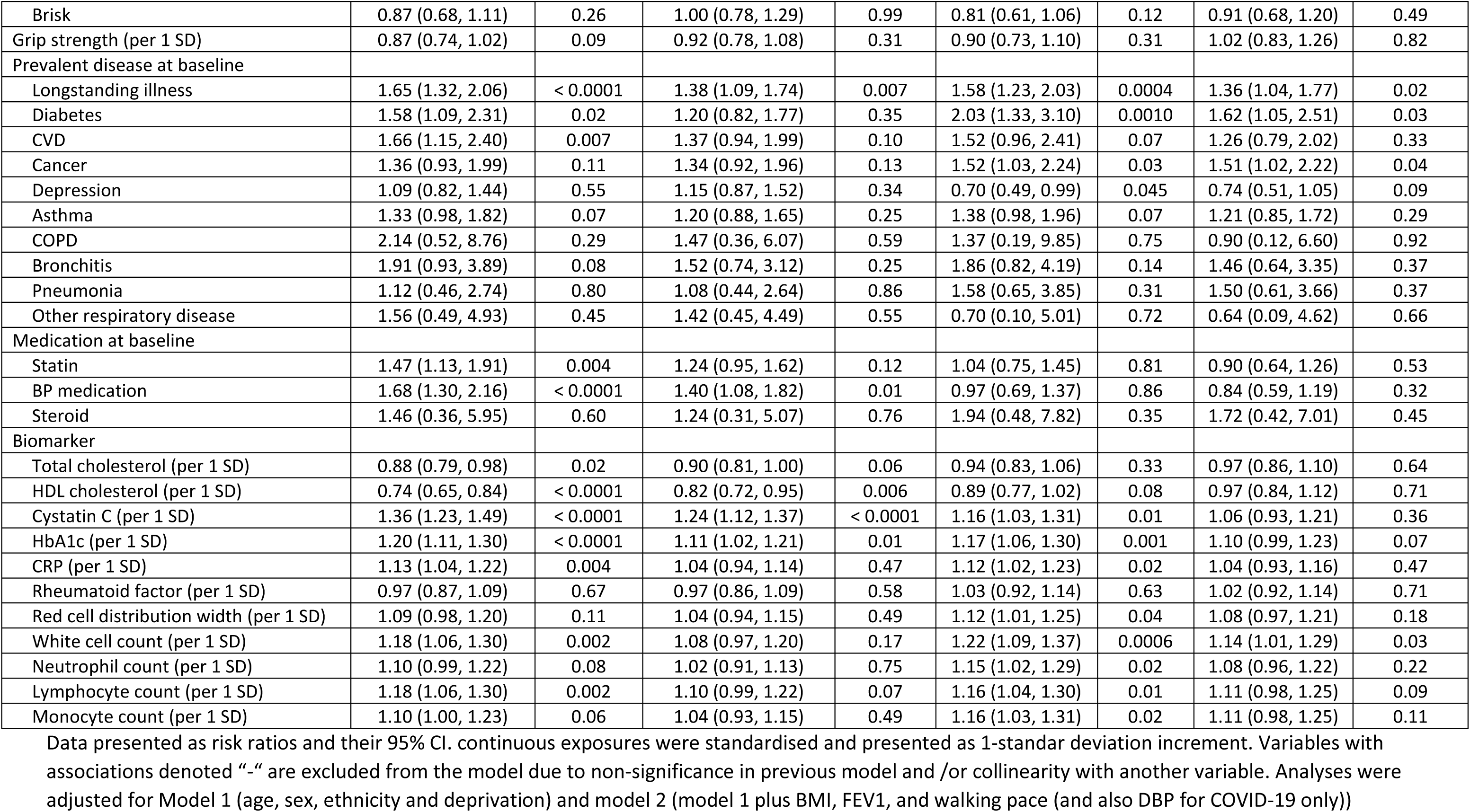
Association of risk factors for COVID-19 and pneumonia in UK Biobank

After adding BMI, blood pressure, FEV1 and walking pace as covariates, modifiable risk factors for COVID-19 admission continued to include smoking (RR 1.38 (95% CI 1.11-1.73)), higher BMI (RR 1.24 (95% CI 1.12-1.38)), use of blood pressure lowering medication as a proxy for hypertension (RR 1.40 95% CI 1.08-1.82) and slow walking pace (RR 1.66 (95% CI 1.20-2.30)). Other risk factors included older age (RR 1.10 per 5 year increase (95% CI 1.02-1.36)), male sex (RR 1.64 (95% CI 1.25-2.14)), black ethnicity (RR 1.86 (95% CI 1.03, 3.36)), socioeconomic deprivation (RR 1.26 (95% CI 1.14-1.40)), longstanding illness (RR 1.38 (95% CI 1.09-1.74)), lower HDL-cholesterol (RR 0.82 (95% CI 1.72-0.95)) and higher cystatin C (RR 1. 24 (95% CI 1.12-1.37)) (Table 2).

Nonlinear associations of continuous variables with COVID-19 and pneumonia are shown in Figure 1 (and supplementary table 2). Age was associated with COVID-19 admission curvilinearly, where participants aged 40-60 years shared similar risk and participants aged over 60 years had exponentially increased risk with age. Associations were similar when adjusting for body fat percentage rather than BMI (Supplementary Table 3).

**Figure 1.**
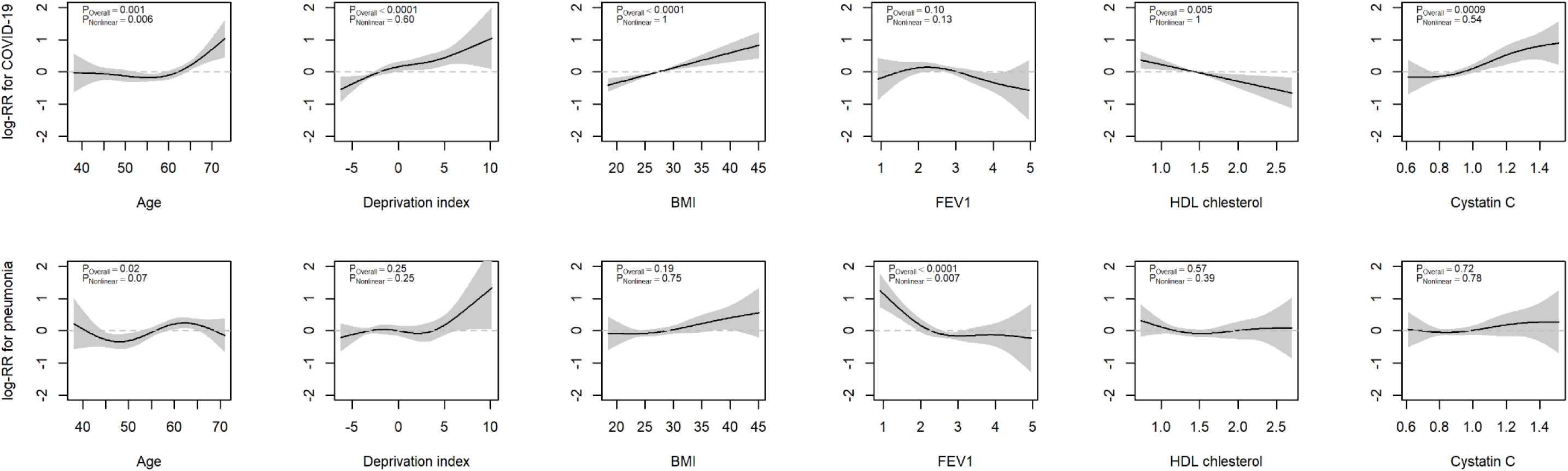
Nonlinear associations of significant continuous variables with COVID-19 and pneumonia

Directly contrasting model 2 for COVID-19 and pneumonia (Figure 2), the risk factors in common for both conditions were older age, higher BMI and slow walking pace, although the latter two associations less strongly associated with pneumonia than for COVID-19. Highlighting specific differences, smoking and treated blood pressure were only associated with COVID-19. Pneumonia was more common in women, and socioeconomic deprivation and cystatin-C were not associated with pneumonia. Low FEV1 was independently associated with pneumonia, but not with COVID-19. Pneumonia also showed an association with baseline cancer.

**Figure 2.**
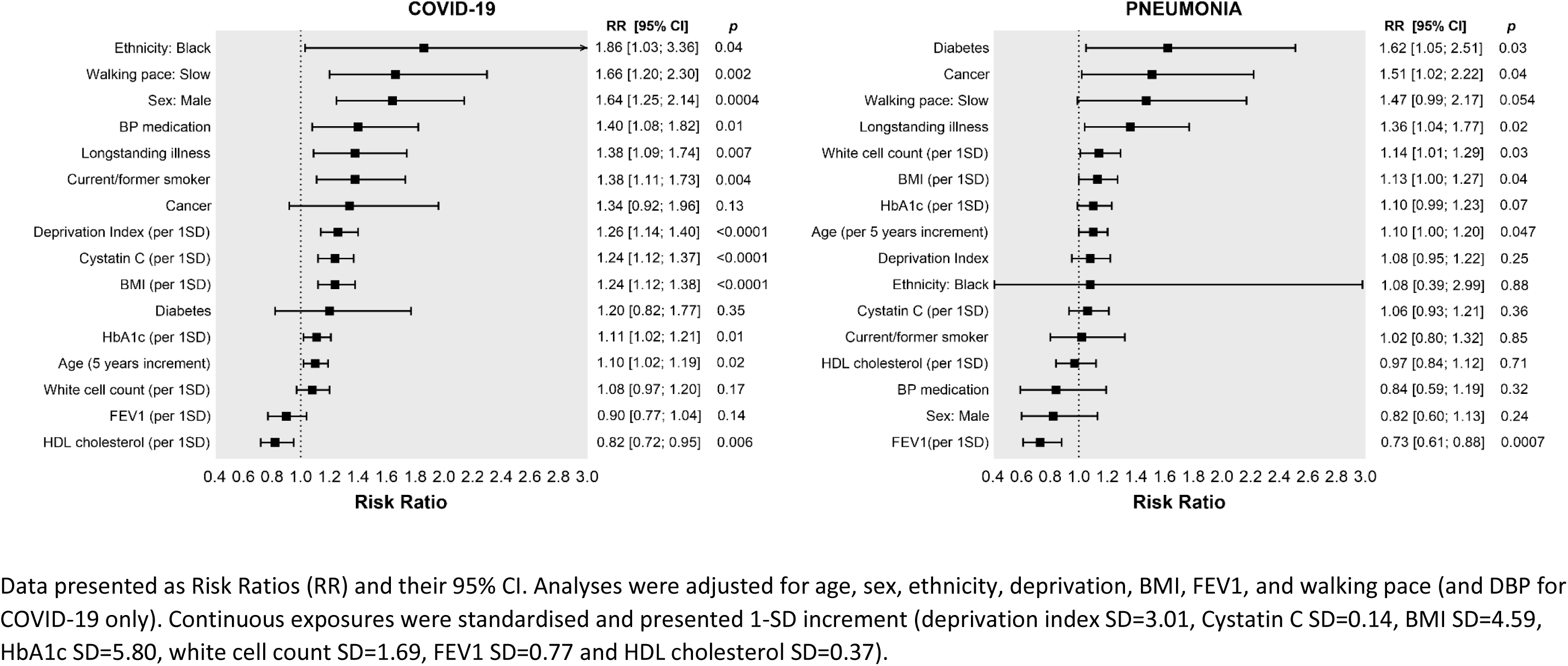
Risk factors for COVID-19 and Pneumonia in the UK Biobank cohort.

### Population attributable risks

Factors that were significant in Table 2 were mutually adjusted to compare their population attributable fractions for COVID-19 and pneumonia (Table 3). Among potentially modifiable risk factors smoking accounted for 12.8% of COVID-19 cases that occurred within the UK Biobank population, treated BP with 5.0% of cases, slow walking pace with 2.8%, and BMI with 0.6% (total 21.2%). In contrast, none of these factors were large contributors to pneumonia cases within UK Biobank. (Table 3)

**Table 3.** Population attributable fractions of COVID-19 in the UK Biobank

|  | COVID-19 |  | Pneumonia after 2016 |  |
| --- | --- | --- | --- | --- |
|  | RR (95% CI) | PAF | RR (95% CI) | PAF |
| Age | 1.04 (0.95 to 1.13) | 33.8% | 1.10 (0.99 to 1.21) | 59.0% |
| Current/former smoker | 1.33 (1.06 to 1.67) | 12.8% | 0.98 (0.76 to 1.27) | -0.8% |
| Male | 1.27 (0.94 to 1.72) | 11.2% | 0.81 (0.57 to 1.15) | -9.5% |
| Longstanding illness | 1.27 (1.00 to 1.62) | 7.3% | 1.28 (0.97 to 1.69) | 7.5% |
| BP medication | 1.33 (1.02 to 1.73) | 5.0% | 0.79 (0.55 to 1.12) | -3.6% |
| Walking pace: Slow | 1.43 (1.02 to 1.99) | 2.8% | 1.27 (0.85 to 1.90) | 1.9% |
| Deprivation index | 1.23 (1.11 to 1.36) | 2.2% | 1.04 (0.92 to 1.19) | 0.2% |
| Baseline cancer | 1.28 (0.88 to 1.87) | 1.9% | 1.47 (0.99 to 2.17) | 3.1% |
| Cystatin C | 1.18 (1.06 to 1.31) | 1.4% | 1.04 (0.92 to 1.19) | 0.1% |
| Ethnicity: Black | 1.89 (1.01 to 3.53) | 1.3% | 1.16 (0.41 to 3.27) | 0.4% |
| HDL cholesterol | 0.87 (0.76 to 1.00) | 0.9% | 1.01 (0.87 to 1.17) | 0.0% |
| HbA1c | 1.12 (1.00 to 1.26) | 0.7% | 1.02 (0.89 to 1.17) | 0.0% |
| BMI | 1.11 (0.99 to 1.25) | 0.6% | 1.06 (0.93 to 1.21) | 0.3% |
| FEV1 | 0.94 (0.81 to 1.09) | 0.2% | 0.77 (0.64 to 0.93) | 3.5% |
| White cell count | 1.01 (0.91 to 1.12) | 0.0% | 1.12 (0.99 to 1.27) | 0.8% |
| Baseline diabetes | 0.70 (0.42 to 1.17) | -1.5% | 1.39 (0.78 to 2.47) | 1.8% |

Estimated using prevalence in the study sample and RR shown in this table.
All factors shown are mutually adjusted.
RR shown in this table may be overadjusted, e.g. cystatin C and HDL cholesterol may be downstream factors of BMI.

## Discussion

In this population-based study, we found that confirmed COVID-19 infection was associated with a number of modifiable risk factors, a trend that less apparent for pneumonia and influenza. In particular, the associations of smoking, BMI (and body fat), hypertension, and physical fitness (as measured by slow walking pace) with COVID-19 are of note; even when such factors were measured a decade before infection they potentially accounted for one-fifth of UK Biobank confirmed COVID-19 cases. The other independent risk factors for COVID-19 infection included older age, male sex, black ethnicity, socioeconomic deprivation, longstanding illness, and reduced renal function as measured by cystatin C, the latter also notable given renal complication in severe COVID-19.

It is importance to recognise the competing risk for health of lockdown and social distancing. Social distancing reduces viral transmission, but also has consequences for lifestyle. Previous data suggests that physical fitness can be rapidly lost when activity levels decrease ^20,21^, and this will also result in an increase in BMI ^22,23^. Further, social distancing may increase loneliness, depression, and psychological stress in some people and consequently adversely affect eating habits ^24^ and other health behaviours. Anecdotal evidence from our clinics and media suggest many are struggling with overeating. This study strongly suggests public health guidance should focus on reducing the risk of severe complications of COVID-19 by advocating a healthy lifestyle during the ongoing pandemic, not just for general cardiovascular and metabolic health, but also to help to protect against COVID-19 infection, used alongside other public health interventions.

Understanding the actual causes of disease, rather than markers that simply correlate with exposures, is clearly a key issue to consider. The independent association of socioeconomic deprivation with COVID-19 after multiple adjustment may be explained by the accumulation of earlier life socioeconomic adversities which can result in less physiological reserve and more multimorbidity ^24,25^. It may also be linked to more overcrowding, reduced social distancing, and potential exposure to greater viral load. Asthma, diabetes and high blood pressure, which have been shown to be associated with a higher risk of severe COVID-19 outcomes ^26^ also showed some trends to be associated with COVID-19 in this dataset. While substantial focus of COVID-19 research has been its apparently more aggressive symptoms and disease progression in older people, it is important to recognise that people who are older have less cardiorespiratory reserve to cope with COVID-19 infection. Older age is also associated with more hypertension and diabetes, poorer lung function, and greater relative fat mass ^24^.

Previous reports have suggested that obesity or excess ectopic fat deposition may be a unifying risk factor for severe Covid-19 infection ^27,28^, reducing both protective cardiorespiratory reserve as well as potentiating the immune dysregulation that appears, at least in part, to mediate the progression to critical illness in a proportion of COVID-19 patients. Our analysis bears out these hypotheses. Once BMI or body fat was adjusted for, inflammatory markers were no longer associated with incident COVID-19. This suggests proinflammatory markers, arising from increased adipose deposition, are probably acting as a marker for body fat which seems to be an adverse risk factor for severe COVID-19 ^27–30^. Furthermore, obesity enhances thrombosis ^31^, which is relevant given the association between severe COVID-19 and pro-thrombotic disseminated intravascular coagulation and high rates of venous thromboembolism ^32,33^, as well as the association with D-dimer seen in other reports ^34^. D-dimer was not measured in blood samples in UK Biobank.

Given that pneumonia is a critical clinical complication of COVID-19, it is important to understand how risk factors for COVID-19 related pneumonia differ from “classic” community acquired pneumonia ^11^. Our comparison between other common respiratory diseases and COVID-19 suggests that identification of at-risk groups based on our understanding of other respiratory diseases may be inadequate - a more refined approach to risk stratification based on COVID-19 specific risks is needed, and future data should focus on some of the exposures we identify to achieve this.

The strengths of our study include the large cohort size at an age relevant to more severe COVID-19 symptoms, and biochemistry assays performed in a single dedicated central laboratory. We were also able to extensively explore emerging and novel risk factors for COVID-19, while simultaneously comparing to risk factors for pneumonia identified to primary care providers. Limitations include that UK Biobank is not representative of the whole UK population,^35^ (our data focuses on England specifically) although this is generally not a concern in investigating risk associations^36^. Care should be taken in generalising the PAF estimates. These related to cases occurring within the UK Biobank population, but are not directly applicable to the general population where the prevalence of risk factors is different. However, they allowed a direct comparison between COVID-19 and pneumonia in the same cohort. Due to the under-representation of non-white ethnicities in UK Biobank, we have limited power to explore important interactions by ethnic group (although Black ethnicity was associated with the outcome), and we recognise this as an important risk factor. Ascertainment bias, including differential healthcare seeking, differential testing and differential prognosis may explain some differences in outcomes given poor coverage of testing in the UK. It is also likely that cases will generally be at the more severe end of the clinical spectrum by using hospitalised cases as the outcome, although use of admission to ITU would be additionally informative once sufficient case numbers accrue. Despite this, we still observe many similar risk factors to incident pneumonia, which is more likely to have close to complete case ascertainment in primary care records, although, as discussed, other risk factors seem more strongly linked to COVID-19 infections. Exposures were measured several years before the development of the outcomes, and misclassification of the exposures will likely bias our results to the null. However, this also serves to illustrate that the risk factors were generally present many years before development of the disease, and as is well known, risk factors tend to track with age. We have also been unable to fully exclude all deaths that occurred prior to the pandemic, due to lack of up-to-date linkage to mortality records.

In conclusion, these early data from UK Biobank suggest risk factors for confirmed COVID-19 infection differ in some important ways from risk factors for pneumonia, being more common in males than females, in lower SES, and with stronger associations with ethnicity, CV risk markers, prior smoking and adiposity. Such findings suggest possible merit in advocating improvements in lifestyle as an additional measure to reduce the risk of COVID-19 alongside existing public health measures such as social distancing and shielding of high risk groups. have implications for health advice targeted at the public to lessen risks during this pandemic

## Data Availability

UK Biobank data can be requested by bona fide researchers for approved projects, including replication, through https://www.ukbiobank.ac.uk/

## Contributors

PW, JPP and NS conceived the idea for the paper. FH conducted the analysis. All authors contributed to the interpretation of the findings. PW, FH, CCM and NS jointly wrote the first draft. All authors critically revised the paper for intellectual content and approved the final version of the manuscript.

## Declarations of interest

JPP is a member of the UK Biobank Steering Committee. Apart from the funding acknowledged below, we declare no other competing interests.

## Acknowledgements

The work in this study is supported by the British Heart Foundation Centre of Research Excellence Grant RE/18/6/34217. CLN acknowledges funding from a Medical Research Council Fellowship (MR/R024774/1). SVK acknowledge funding from the Medical Research Council (MC_UU_12017/13) and Scottish Government Chief Scientist Office (SPHSU13). SVK also acknowledges funding from NRS Senior Clinical Fellowship (SCAF/15/02).

